# The Role of Competency-Based Equipment User Training in Enhancing Healthcare Providers’ Skills in Utilizing bCPAP Devices for Newborn Care in Dar es Salaam, Tanzania: A Qualitative Study

**DOI:** 10.64898/2026.09.25.26363989

**Authors:** Josephat Mutakyamilwa, Alen Kinyina, Robert Tillya, Elizabeth Ngowi, Hope Peter, Josephine Shabani, Donat Shamba, Honorati Masanja, Mariam Thabit Johari, Nahya Salim

## Abstract

**Introduction:** Bubble Continuous Positive Airway Pressure (bCPAP) is an effective intervention for managing respiratory distress syndrome among preterm newborns. However, its successful utilization depends on healthcare providers possessing adequate knowledge and practical skills. In Tanzania, competency-based equipment user training has been introduced to strengthen providers’ capacity to use neonatal technologies, including bCPAP. Despite these efforts, there is limited qualitative evidence on how this training influences healthcare providers’ competence and routine clinical practice. This study explored the role of competency-based equipment user training in enhancing healthcare providers’ skills in utilizing bCPAP devices for newborn care in Dar es Salaam, Tanzania.

**Methods:** A phenomenological qualitative study was conducted in three regional referral hospitals implementing the NEST360 programme. Purposive sampling was used to recruit doctors and nurses who had completed competency-based equipment user training. Data were collected through in-depth interviews using a semi-structured interview guide and analysed using Braun and Clarke’s thematic analysis approach.

**Findings:** Six major themes emerged. Participants described competency-based training as a great approach for improving practical skills, confidence, and clinical decision-making regarding patient selection, device operation, monitoring, and weaning. Participants also reported improved equipment maintenance practices, reduced preventable equipment damage, and stronger collaboration with biomedical engineering professional. The training promoted positive changes in provider behaviour, including greater autonomy, internal mentorship of newly recruited staff, and increased accountability for equipment management. Participants further perceived improvements in neonatal care through earlier initiation of bCPAP, fewer unnecessary referrals, and better management of preterm newborns within their facilities.

**Conclusion:** Competency-based equipment user training strengthened healthcare providers’ competence and confidence in utilizing bCPAP for newborn care while improving equipment management and interdisciplinary collaboration. Integrating competency-based training into routine newborn care programmes may enhance the effective implementation and sustainability of life-saving neonatal technologies in resource-limited settings.

## Introduction

Preterm birth complications account for the largest proportion of neonatal deaths globally. Each year around 15 million newborns are born before 37 complete weeks of gestation[1]. Neonatal deaths among premature babies contribute approximately 33% of 2.3 million deaths occurring each year[2]. In 2024, neonatal deaths contributed approximately 47 per cent of all under-five deaths globally[3]. The World Health Organization (WHO) estimates over 80% of these deaths occur in Sub-Saharan Africa (SSA) and Southern Asia[2,3]. In SSA, neonatal mortality remains high at 27 deaths per 1,000 live births, which is more than double the Sustainable Development Goal (SDG) target aiming to reduce neonatal mortality to at least 12 deaths per 1,000 live births by 2030[2,3].

In Tanzania, approximately 336,000 babies are born prematurely each year, which translates to about 17% of all live births[4]. Prematurity alone contributes almost one third of neonatal mortality followed by birth asphyxia (33%) and infections (24%)[5,6]. To reduce the current NMR by half, Every Newborn Action Plan (ENAP) recommends that at least 80% of districts or equivalent facilities have a functional neonatal care unit (NCU) and use continuous positive airway pressure (CPAP) devices for respiratory support[7,8]. However, a baseline cross-sectional study conducted by Bundala et al.[8] reported that only 31% of health facilities providing comprehensive emergency obstetric and newborn care services in Tanzania had a functioning NCU. It also highlighted that only 3% of these facilities had CPAP devices for managing respiratory distress syndrome (RDS)[8].

Respiratory distress syndrome is a major life-threatening complication of prematurity. One of the prospective studies conducted in Tanzania reported 31.3% deaths among all neonates with RDS[9]. The clinical signs of RDS manifest within the first minutes or hours after birth. RDS results from surfactant deficiency in immature lungs, leading to alveolar collapse and impaired gas exchange. In turn, this can rapidly progress to life-threatening hypoxia if not appropriately managed[10]. In resource-limited settings, the burden of RDS is exacerbated by inadequate respiratory support and limited availability of skilled personnel[9,11]. Bubble CPAP (bCPAP) has been widely recognized as an effective, low-cost intervention for managing RDS[12,13].

In 2023, a randomized trial was conducted in Tanzania to compare the treatment outcomes of bCPAP versus oxygen therapy among preterm babies presenting with respiratory distress. This study reported that babies in the bCPAP group had higher survival (77.3%) as compared to their counterparts in the oxygen therapy group (47.8%)[14]. Furthermore, neonates treated with bCPAP had 52% lower risk of death compared to neonates receiving oxygen therapy[14]. Bubble CPAP significantly reduces mortality among preterm neonates by maintaining airway patency, improving oxygenation, and decreasing the need for invasive ventilation[12,14]. In some settings, bCPAP has been reported to be effective in treating RDS in preterm babies with varying levels of effectiveness ranging from 42% to 85%[14]. Another study conducted at Muhimbili National Hospital reported that bCPAP is feasible to implement due to easy device operation, low resource utilization, and low maintenance requirements[13].

The effectiveness of CPAP is highly dependent on correct device setup, patient monitoring, and timely clinical decision-making, all of which require a competent and well-trained healthcare workforce[11,12,14]. To address some of the challenges in Tanzania, the Newborn Essential Solutions and Technologies (NEST360) alliance in collaboration with the government, has introduced competency-based training approaches aimed at strengthening healthcare providers’ skills in neonatal care, including CPAP use[15]. Unlike traditional didactic training models, competency-based training emphasizes practical skill acquisition, simulation-based learning, continuous assessment, and mentorship to ensure that providers can effectively apply knowledge in clinical settings[16]. These approaches align with the national and global recommendations advocating for skills-based training to improve the quality of care for small and sick newborns.

Despite these efforts, there remains limited qualitative evidence exploring how competency-based equipment user training specifically influences healthcare providers’ skills, experiences, and day-to-day utilization of CPAP devices in real-world clinical settings in Tanzania. Existing studies have largely focused on clinical outcomes and effectiveness of devices[14] or availability of the bCPAP technology[8]. Other studies focused on feasibility and acceptability of device utilization or knowledge of the healthcare providers on usage of the CPAP[11,13]. Variability in trainings (clinical competence), users’ knowledge and contextual factors across facilities may affect the utilization and effectiveness of bCPAP[11]. This gap underscores the need for in-depth qualitative inquiry to better understand how competency-based training shapes provider competence and utilization of CPAP technology. Therefore, this study aimed to explore the role of competency-based equipment user training in enhancing healthcare providers’ skills in utilizing bCPAP devices for newborn care.

By generating an in-depth understanding of healthcare providers’ experiences with competency-based training and bCPAP utilization, the study provides evidence-based knowledge for the development of more effective and context-specific training models that address existing skill gaps and implementation barriers. The results may guide policymakers and stakeholders in optimizing investments in neonatal care technologies like bCPAP by ensuring that training approaches are aligned with real-world clinical needs. Additionally, this study will contribute to the growing body of evidence on health workforce capacity strengthening, particularly in relation to competency-based approaches for utilization of complex medical devices.

## Methodology

### Study design

This study utilized a phenomenological qualitative design to explore the impact of competency-based equipment user training on healthcare providers’ ability to use bubble CPAP (bCPAP) devices in newborn care. Through this design, the study aimed to uncover how various components of the training such as the competency-based curriculum, training materials, duration, and delivery methods shaped participants’ skills and confidence in applying bCPAP in clinical settings.

### Study area

This study was conducted in the Dar es Salaam region, Tanzania. Specifically within three healthcare facilities implementing the NEST360 program: Temeke, Amana, and Mwananyamala Regional Referral Hospitals. These sites were selected because a total of 115 healthcare providers from these hospitals had participated in a structured, four-day, hands-on training program focused on the proper use of newborn care equipment, including bCPAP devices.

NEST360 is an alliance of 23 organizations, 18 of which are based in Africa, working with governments in five African countries, including Tanzania, to meet the SDG 3.2 goal of improving newborn survival. The program provides a comprehensive health systems package, which includes a bundle of technologies to deliver life-saving care for small and sick newborns (SSNC) in hospitals. A central aspect of NEST360 is competency-based training for healthcare providers, including nurses, midwives, doctors, and biomedical engineering professionals, with a focus on the correct use, maintenance, and troubleshooting of essential medical devices, such as the bCPAP, to improve newborn survival rates. The program also focuses on improving data systems to ensure data for action in quality improvement strategies

### Study population and intervention description

The study population comprised healthcare providers who participated in competency-based equipment user training at three NEST360-implementing facilities in Dar es Salaam. These included doctors, nurses, and biomedical engineering professionals. To build competency and ensure safe equipment use, equipment user training followed a three-stages including, preparation, training and post training support (Fig 1).

**Fig 1:**
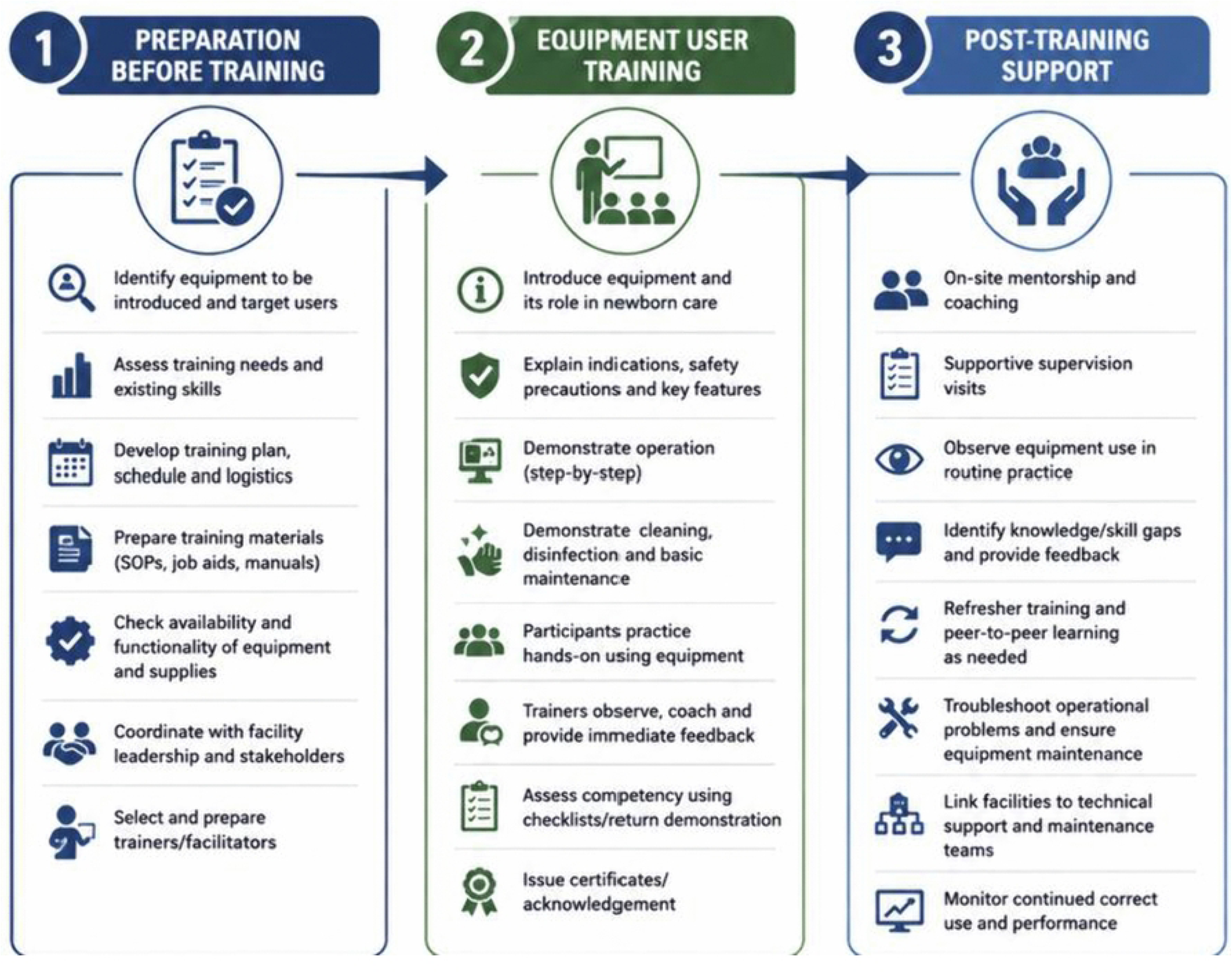
Intervention framework.

### Sample size and selection

Participants were selected using a purposive sampling approach to ensure representation across professional cadres and varying levels of experience. From each of the three hospitals, two enrolled nurse, two registered nurses, two general medical doctors and two pediatrician were included making a total of 24 participants who completed the training and were selected for participation in in-depth interviews. This sampling strategy allowed for the inclusion of diverse perspectives, reflecting differences in roles, educational background, and clinical experience in neonatal care.

### Data collection methods

Data was collected using in-depth interviews (IDIs) guided by a semi-structured interview tool. The guide was designed to explore participants’ experiences with bubble CPAP use prior to the training, how the competency-based equipment user training influenced their utilization of the device in newborn care, and their self-perceived competency in equipment use, including maintenance and minor troubleshooting.

The interview guide was initially developed in English and then translated into Kiswahili to ensure clarity and cultural relevance for participants. This approach minimized language barriers and supported more open and authentic responses. The interview guide was piloted with nurses and doctors who attended the training on the use of bubble CPAP, and piloted data obtained was used to refine paraphrasing of questions and was not included in analysis.

### Data collection procedure

Data collection was conducted over a four-week period by a trained research assistant who is a registered nurse with experience in neonatal care. The assistant underwent a three-day training covering, study objectives and methodology, familiarization with the interview guide, ethical considerations, confidentiality, interview techniques, communication skills, and awareness of interviewer bias. Before the start of data collection permission and approval to collect data at the facility were obtained from the medical officer incharge.

Between 3rd and 17th March 2025, healthcare providers participated in face-to-face interviews arranged via phone calls. Each interview lasted 35 to 45 minutes and was conducted in a private setting within the health facilities to ensure confidentiality. Upon arrival at each neonatal unit, the research team presented an official approval letter to the unit in-charge and the participants as evidence of institutional permission and awareness of the study.

Prior to the interviews, participants were provided with a written informed consent form to allow adequate time for review and consideration of the study’s purpose, procedures, and their rights as participants. Once participants gave written informed consent, the interviews were conducted using a semi-structured interview guide, which explored their experiences with bubble CPAP use prior to the training, the impact of the competency-based training on their clinical practice, and their perceived competency in using, maintaining, and troubleshooting the equipment post-training.

All interviews were conducted in Kiswahili and audio-recorded, with field notes taken to capture non-verbal cues, participant emotions, and contextual details not fully reflected in the recordings. These notes supplemented the transcripts, aided interpretation, and supported daily debriefing sessions between the principal investigator (PI) and the research assistant, ensuring data quality, completeness, and timely resolution of any emerging issues.

### Data analysis

Qualitative data analysis for this study employed Braun and Clarke thematic analysis[17,18]. Initially, the collected data underwent verbatim transcription done by the research assistant immediately after the data collection day. Two researchers (JM & AK) reviewed the transcripts against the audio-recordings to ensure their quality and accuracy before they were imported into qualitative data management software (Nvivo v.12) for analysis.

To avoid loss or distortion of important details and expressions, which were essential for understanding participants’ experiences and perception of using CPAP before the training and after training. Through repeated reading of the transcripts, the researchers familiarized with the data to identify preliminary themes and carefully chose quotes that effectively illustrate various perspectives and patterns relevant to the study objectives. The codes were categorized into themes that link the research questions and data representing a patterned meaning within data. Throughout the analytical process, conceptualization of the keywords, codes and themes was done, and any newly identified codes were incorporated. Any discrepancy was resolved through joint discussions. Findings were interpreted within thematic areas, and key lessons learned were documented. The reporting phase involved the removal of repetitions, filler words, and hesitations, as they contribute no substantive value to the context and translations to English language was done on the quotations included in the report. Ultimately, the analysis culminated in the presentation of key themes and ideas derived from the study.

### Ethical considerations

Ethical clearance for this study was granted by the Muhimbili University of Health and Allied Sciences (MUHAS) Research and Ethics Review Committee referral (REC) Ref.No.DA.282/298/01.C/2531 Following approval, an official introduction letter was issued by the Director of Postgraduate Studies. Additional permissions were obtained from the Medical Officers In-Charge at Amana, Mwananyamala, and Temeke Regional Referral Hospitals, the health facilities where data collection was conducted. At each facility, the research team presented the study objectives, methodology, and potential benefits to ensure institutional understanding and support.

Prior to data collection, written informed consent was obtained from all participants. Each participant was clearly informed about the purpose of the study, procedures involved, their right to voluntary participation, and the option to withdraw at any stage without any consequence. To ensure confidentiality, interviews were conducted in private settings chosen by the participants. Participants were identified using unique codes rather than names, and all personal data were kept strictly confidential and accessible only to the research team.

## Results

### Characteristics of the participants

A total of 24 healthcare providers participated in the in-depth interviews (IDIs). The participants were equally distributed across four professional cadres, comprising pediatricians (n = 6, 25.0%), general medical doctors (n = 6, 25.0%), registered nurses (n = 6, 25.0%), and enrolled nurses (n = 6, 25.0%). Regarding professional experience, 11 (45.8%) participants had more than five years of work experience, 7 (29.2%) had between two and five years, and 6 (25.0%) had less than two years of experience. In terms of educational attainment, 6 (25.0%) held certificate-level qualifications, and 4 (16.7%) had diploma-level qualifications, 8 (33.3%) participants held bachelor’s degrees **and** 6 (25.0%) had master’s degrees,. Participants were recruited equally from the three study sites, with 8 (33.3%) participants each from Amana, Mwananyamala, and Temeke Regional Referral Hospital.

**Table 1.** Characteristics of the participants.

| Characteristic | Category | n (%) |
| --- | --- | --- |
| Participant category | In-depth interview (IDI) participants | 24 (100.0) |
| Professional cadre (IDIs, n = 24) | Pediatricians | 6 (25.0) |
|  | General medical doctors | 6 (25.0) |
|  | Registered nurses | 6 (25.0) |
|  | Enrolled nurses | 6 (25.0) |
| Work experience (IDIs, n = 24) | < 2 years | 6 (25.0) |
|  | 2–5 years | 7 (29.2) |
|  | > 5 years | 11 (45.8) |
| Level of education (IDIs, n = 24) | Master's degree | 6 (25.0) |
|  | Bachelor's degree | 8 (33.3) |
|  | Diploma | 4 (16.7) |
|  | Certificate | 6 (25.0) |
| Health facility (IDIs, n = 24) | Hospital A | 8 (33.3) |
|  | Hospital B | 8 (33.3) |
|  | Hospital C | 8 (33.3) |

### Theme 1: Experience using CPAP before the training

Prior to the introduction of competency-based training, Health Care Providers in Dar es Salaam reported to have encountered critical barriers in effectively utilizing bCPAP for neonatal care. Despite the physical availability of the bCPAP machines in some facilities, most providers lacked theoretical knowledge and practical skills. Providers commonly lacked knowledge about criteria for starting or weaning off CPAP. This deficit in procedural competence created ethical and emotional dilemmas for staff. For instance, a paediatrician working in one of the high-volume hospitals said:

> *“Before receiving training, we didn’t know how to use them properly. We were given machines, but we weren’t provided with thorough training…. So, we often didn’t know which child was eligible for CPAP, nor how to monitor a baby while on the device. We didn’t even know how to detect minor faults in the equipment, like identifying when the machine wasn’t functioning or had completely failed.”* (Paediatrician, Hospital B).

Another nurse added

> "*Before the training, I didn’t know where to begin or what to monitor…. I would just start using the device without understanding it, and if I saw it bubbling or heard a sound, I would assume it was working. I had no idea what to check to identify if the CPAP was not working. Sometimes we thought the machines were cheap or low quality, but in reality, we just didn’t know how to use them properly."* (Nurse, Hospital B)

### Theme 2: Improved practical skills and confidence of healthcare providers

The introduction of competency-based training enhanced healthcare providers’ knowledge and confidence in using bCPAP machines for neonatal care. Before the training, even experienced healthcare workers operated the equipment with limited comprehension of its clinical indications or monitoring requirements. The training empowered them to make informed decisions, interpret clinical cues, and use CPAP both for treatment and as a preventive intervention. As one general doctor from Hospital A explained:

> *“The training really opened our eyes. We learned when CPAP should be used, how to identify which babies could benefit from it, and how to assemble it correctly… We also learned how to clean and maintain the machine after use. This knowledge made my work easier and gave me the confidence to face challenges.” (General Doctor, Hospital A)*.

Other participants emphasized that training not only enhanced technical skills but also contributed to facility-level autonomy and reduced dependence on higher-level referral centers.

> *“We learned that even a baby under 32 weeks without signs of RDS can benefit from CPAP as a preventive treatment. From that point on, we started managing such cases ourselves instead of referring them to Muhimbili.” (*Paediatrician, Hospital A).

Another participant added that:

> *"Now we train new staff and conduct internal mentorship. Even when only a few staff who received formal training remain in the neonatal unit, we continue to use ward rounds as an opportunity to teach others. Over time, we have trained many new staff members and interns, helping to sustain and strengthen the use of CPAP in our unit."* (Registered Nurse, Hospital A).

### Theme 3: Reduced equipment damage and improved maintenance

Participants described substantial changes in equipment handling and maintenance practices following competency-based training on neonatal CPAP. Prior to the training, healthcare providers reported frequent equipment breakdowns, limited maintenance knowledge, and shortages of essential consumables that often-necessitated improvisation to keep CPAP machines operational. One participant explained:

> *“Before training, many devices were breaking down—not just CPAPs. We had two CPAP machines, and both were non-functional. We didn’t have enough tubing, and sometimes we would cut catheters and join them to make tubes.”* (General Medical Doctor, Hospital C)

Participants indicated that such practices reflected limited technical capacity to maintain equipment alongside inadequate availability of essential supplies.

Following the training, participants reported improved knowledge and skills in the operation, handling, and maintenance of CPAP machines. They described increased confidence in conducting routine equipment checks, identifying and resolving minor technical problems, distinguishing reusable from disposable components, and collaborating with biomedical technicians to prevent equipment failure. As one participant stated:

> *“Thanks to the training, we now use the machines correctly, perform routine checks, and collaborate with technicians on preventive maintenance. We can replace water, check pressure, inspect for leaks, and even handle minor repairs ourselves. This has greatly reduced breakdowns, especially at night when technicians aren’t available, allowing us to keep machines running until help arrives.”* (Enrolled Nurse, Hospital C)

Other participants noted that after the training, some facilities institutionalized protocols and systems to support equipment longevity. Dedicated cleaning areas were established, protocols for daily and periodic checks were introduced, and collaboration with biomedical engineers was strengthened. For instance, a nurse said:

> *“We established a clear protocol for cleaning the equipment and designated a specific area for that purpose. The hospital has also hired biomedical engineers who assist us regularly, and periodic preventive maintenance (PPM) is now being done”* (Enrolled Nurse, Hospital C).

### Theme 4: Improved healthcare provider behaviour

One of the most impactful outcomes of the competency-based CPAP training was a noticeable shift in the behaviour, clinical decision-making, and care culture among healthcare providers. Before the training, CPAP use was characterized by hesitation, over-reliance on physician supervision, and delayed initiation of therapy. Following the training, participants adopted a more proactive and confident approach to CPAP management. Providers began initiating therapy earlier, based on clinical signs, and demonstrated autonomy in adjusting settings, monitoring response, and decreasing and weaning CPAP treatment appropriately. A paediatrician from Hospital A illustrated how the training directly influenced referral patterns and strengthened facility-based care:

> *“Before training, we lacked confidence and frequently referred babies to high level hospital [he named it] often calling an ambulance daily…Now, we manage preterm babies ourselves, reducing referrals, ambulance use, and hospital costs. Thanks to the training, we can go three to four days without a single referral, a significant improvement from before* (Paediatrician, Hospital A).

Another Nurse added that

> *“Before training, I didn’t even know what FiO₂ [parameter] was. Now I can increase or decrease oxygen levels based on the baby’s condition, and I know when it’s time to start or wean off CPA.*” (Registered Nurse, Hospital B).

Other participants highlighted that the training reinforced a culture of accountability and respect for equipment. Many participants began to take ownership of maintaining CPAP devices, ensuring cleanliness and proper decontamination, and promoting proper storage practices.

> *“Now I understand the importance of each piece of equipment. If I see items stored carelessly or left out, I immediately return them to a safe place to avoid misplacing them”* (Registered Nurse, Hospital A).

Another Nurse said:

> *“This training has greatly improved my daily work, boosting my confidence in the NICU. I now operate the equipment more competently, troubleshoot minor issues, and communicate effectively with biomedical staff”* (Enrolled Nurse, Hospital C).

### Theme 5: Impact on newborn outcomes and unit efficiency

One of the most significant and encouraging results of the competency-based CPAP training was its tangible impact on both newborn health outcomes and the overall efficiency of neonatal units. Participants consistently reported a reduction in neonatal mortality, especially among preterm and low birth weight babies. This improvement was largely attributed to the earlier initiation of CPAP, better clinical decision-making, and timely management of respiratory distress, which had previously led to avoidable deterioration or death.

A general doctor from Hospital A explained how these changes directly translated into better survival outcomes:

> “*With CPAP and proper training, most babies are now treated here instead of being referred. Early care has reduced mortality, unnecessary referrals, and this approach has saved lives, minimized healthcare costs, and eased the financial and emotional burden on families”* (General Doctor, Hospital A).

Participants reflected how the training enabled them to respond more promptly, allowing timely interventions that they perceived as critical in supporting newborns’ survival.

> *“Previously, we waited until the baby showed symptoms… Now, for any baby under 32 weeks, we start CPAP early. This has significantly improved care outcomes and reduced deaths” (*General Doctor, Hospital C).

### Theme 6: Improved collaboration with biomedical engineering professionals

Before the competency-based CPAP training, most neonatal care facilities faced critical gaps in technical support for medical equipment. The training acted as a catalyst in redefining the relationship between clinical staff and biomedical support teams. Healthcare providers gained not only technical knowledge about the machines they used but also a better appreciation of the roles and responsibilities of biomedical technicians. One paediatrician said

> *“After the training, the hospital realized the importance of having a biomedical technician. They even hired one on a temporary contract because services cannot continue without them”* (Paediatrician, Hospital B).

Another participant added

> ”*Now we work hand in hand with biomedical staff, I can clearly report when a particular machine isn’t working and describe the symptoms accurately….and now technicians better understand the clinical impact and respond faster, knowing equipment failure can threaten a baby’s life. This collaboration has improved response times and reduced our dependence on them for minor issues*” (Enrolled Nurse, Hospital C).

Participants noted that the training emphasized the importance of preventive maintenance. Providers learned that routine servicing of CPAP machines was essential to prevent breakdowns and ensure consistent performance. As a result, staff began actively coordinating with biomedical teams to schedule regular checks and upkeep.

> *“We learned the importance of preventive maintenance (PPM) and now collaborate closely with biomedical technicians to ensure it’s done regularly. We remind them when maintenance is due and perform routine maintenance ourselves. This has improved equipment durability and reduced breakdowns significantly”* (Enrolled Nurse, Hospital C).

## Discussion

### Improved healthcare providers’ competence and confidence following competency-based equipment user training

This study demonstrates that competency-based equipment user training substantially improved healthcare providers’ practical competence, confidence, and clinical decision-making regarding the use of bCPAP in newborn care. Prior to the training, participants consistently described limited knowledge regarding patient selection, device assembly, monitoring, troubleshooting, and weaning from CPAP, despite the availability of the equipment in their facilities. These findings suggest that competency-based training addressed a critical implementation gap by transforming the availability of technology into effective clinical utilization.

Our findings are consistent with previous studies conducted in Kenya[21] and Malawi[22], which reported that the successful implementation of CPAP depends not only on equipment availability but also on HCPs possessing adequate technical competence and confidence. A qualitative study from Kenya found that inadequate knowledge and insufficient training were among the greatest barriers preventing safe and consistent CPAP utilization, while HCPs emphasized that structured training substantially improved their confidence and willingness to use the technology[21]. Likewise, an implementation study from Malawi demonstrated that frequent CPAP use was strongly associated with facilities where healthcare providers had received adequate training and accumulated practical experience using the equipment[22]. These findings reinforce the argument that investments in neonatal technologies alone are insufficient unless accompanied by systematic capacity strengthening of frontline healthcare workers.

One notable finding was that participants described competency-based training as fundamentally different from conventional didactic approaches. Rather than merely acquiring theoretical knowledge, providers emphasized learning through hands-on practice, simulation, repeated demonstrations, and supervised clinical application. This observation aligns closely with principles of competency-based medical education, which emphasize demonstration of observable competencies rather than passive knowledge acquisition. Simulation-based neonatal education allows providers to repeatedly practice complex procedures in a controlled environment before applying them in clinical practice, thereby improving procedural competence, confidence, and patient safety[23].

The improvement in providers’ confidence described in this study may also be explained through experiential learning theory[24]. According to this theory, competence develops through repeated cycles of concrete experience, reflective observation, conceptual understanding, and active experimentation. During competency-based training, healthcare providers actively assembled CPAP machines, identified clinical indications, interpreted patient responses, and practiced troubleshooting under supervision. Such experiential learning enables participants to integrate theoretical knowledge with practical application, thereby increasing self-efficacy and facilitating long-term retention of clinical skills. Unlike lecture-based training, competency-based approaches allow immediate feedback and correction of errors, reducing uncertainty when providers subsequently encounter similar clinical situations[25].

An important contribution of this study is the observation that enhanced competence extended beyond individual performance to influence organizational learning. Several participants reported subsequently mentoring newly recruited staff, interns, and colleagues through ward rounds and on-the-job coaching. This suggests that competency-based training generated secondary benefits by strengthening local training capacity and promoting knowledge transfer within neonatal units. Similar findings were reported from Kenya, where a structured training-of-trainers model successfully enabled locally trained providers to transfer CPAP knowledge and practical skills to subsequent generations of healthcare workers without compromising training quality[26]. Such cascading mentorship models may represent a cost-effective strategy for sustaining competency in settings characterized by frequent staff turnover and limited opportunities for external training.

The findings also highlight that competency-based training strengthened providers’ clinical reasoning rather than simply teaching technical procedures. Participants described improved ability to assess eligibility, interpret clinical signs, adjust oxygen concentrations appropriately, monitor treatment response, and determine when CPAP could safely be discontinued. This shift reflects progression from task-oriented equipment operation toward comprehensive clinical decision-making. Previous implementation research has similarly emphasized that successful neonatal technology adoption requires providers to understand the clinical principles underpinning technology use rather than merely learning operational steps[27]. Such higher-order competencies are particularly important in resource-limited settings where providers frequently manage critically ill newborns with limited specialist supervision.

Collectively, these findings suggest that competency-based equipment user training serves as a health systems intervention rather than merely an educational activity. By strengthening healthcare providers’ competence, confidence, clinical autonomy, and mentorship capacity, the training enhances facilities’ ability to deliver high-quality respiratory care for small and sick newborns. These improvements may ultimately facilitate wider adoption of life-saving neonatal technologies while reducing unnecessary referrals to tertiary hospitals.

### Reduced equipment damage and improved equipment maintenance

A second major finding of this study was that competency-based equipment user training substantially improved healthcare providers’ ability to properly use, maintain, preserve, and troubleshoot bCPAP equipment. Before the training, participants frequently reported uncertainty regarding routine maintenance procedures, cleaning protocols, identification of equipment malfunctions, and differentiation between reusable and disposable components. Following the training, providers described greater confidence in performing preventive maintenance, conducting routine equipment checks, identifying minor technical faults, and collaborating effectively with biomedical engineering professionals. These improvements were perceived to reduce equipment downtime and enhance the continuous availability of functional CPAP machines for neonatal care.

This finding is particularly important because equipment failure remains a major barrier to effective neonatal care across many low-resource settings. Previous studies have shown that although CPAP devices may be available in health facilities, inadequate maintenance systems, improper handling, and limited technical capacity often result in frequent equipment breakdowns and interrupted clinical services[22]. In Tanzania, the availability of neonatal technologies has improved substantially in recent years through national investments and programmes such as NEST360, yet maximizing their impact requires parallel investments in healthcare workers’ capacity to operate and maintain these technologies effectively.

The improvements reported in this study may be explained by the practical orientation of competency-based training. Rather than focusing exclusively on clinical indications for CPAP, the training equipped participants with practical knowledge regarding equipment assembly, cleaning, infection prevention, routine inspection, and basic troubleshooting. Consequently, providers developed a stronger sense of ownership over the equipment and became active participants in preserving valuable hospital resources. This behavioural change reflects broader principles of competency-based education, whereby learners acquire integrated competencies encompassing technical skills, professional responsibility, and problem-solving abilities instead of isolated procedural knowledge.

An important observation emerging from this study was the institutionalization of preventive maintenance practices within the neonatal units. Participants described establishing dedicated cleaning areas, implementing standardized maintenance protocols, conducting regular equipment inspections, and working more closely with biomedical engineering professionals to schedule preventive maintenance. These findings suggest that competency-based training generated organizational changes extending beyond individual competence. Rather than simply improving providers’ knowledge, the intervention appeared to influence unit-level practices and foster a culture of equipment stewardship.

These findings are consistent with implementation science perspectives that emphasize technology adoption as a systems process involving interactions between people, organizational structures, and technical infrastructure. Successful implementation depends not only on the technical characteristics of the equipment but also on users’ competencies, communication pathways, maintenance systems, and institutional support[27]. The improvements reported by participants therefore likely reflect enhanced integration of these interacting components rather than the isolated effects of training alone.

From a health systems perspective, reducing preventable equipment damage carries important economic implications. CPAP devices represent valuable investments for hospitals in low-resource settings, where procurement budgets remain constrained and replacement of damaged equipment may take months. By equipping healthcare providers with basic maintenance and troubleshooting skills, competency-based training may reduce repair costs, extend equipment lifespan, and increase the proportion of time that devices remain clinically available. Such efficiency gains are particularly important in settings experiencing high patient volumes, shortages of respiratory support equipment, and limited biomedical engineering capacity.

### Implications for policy, practice, and future research

This study provides important evidence that competency-based equipment user training can substantially strengthen healthcare providers’ competence and confidence in utilizing bubble CPAP for newborn care in resource-limited settings. The findings suggest that strengthening provider competency should be considered an integral component of introducing neonatal technologies rather than a complementary activity. In particular, the study demonstrates that practical, simulation-based training combined with ongoing mentorship equips healthcare providers with the skills required not only to operate CPAP safely but also to perform routine equipment maintenance, troubleshoot minor faults, and make timely clinical decisions. These findings may therefore inform national newborn care programmes and hospital managers in designing implementation strategies that integrate competency-based training into routine neonatal services to improve the quality of care for small and sick newborns.

The findings also provide practical insights into the successful implementation and sustainability of neonatal technologies within routine health systems. The identification of improved collaboration between healthcare providers and biomedical technicians, increased provider confidence, and enhanced equipment stewardship highlights key implementation components that should be prioritized during programme expansion. These findings suggest that scaling up bubble CPAP should extend beyond equipment procurement to include structured competency assessment and training, hands on mentorship & supportive supervision, periodic refresher training, and interdisciplinary collaboration. Such approaches may strengthen equipment functionality, reduce preventable breakdowns, improve continuity of newborn care, and maximize returns on investments in neonatal technologies, particularly in low-resource settings where equipment replacement is often limited. These observations are consistent with recent implementation research demonstrating that sustainable technology adoption depends on strengthening workforce capacity and organizational systems alongside technology introduction.

Finally, this study contributes to the limited implementation science literature examining competency-based equipment user training for neonatal technologies in sub-Saharan Africa. Unlike previous studies that primarily evaluated CPAP effectiveness, feasibility, or provider knowledge, this study provides an in-depth understanding of how competency-based training influences healthcare providers’ clinical practice, equipment management, and interdisciplinary teamwork in routine service delivery. Future research should build upon these findings by examining the long-term sustainability of competency-based training, its impact on objectively measured provider competencies and neonatal outcomes, and the effectiveness of mentorship and training-of-trainers models across different levels of the healthcare system. Such evidence will be important for informing context-specific strategies for scaling neonatal technologies and strengthening newborn care services in Tanzania and similar low-resource settings.

### Strengths of the study

This study has several important strengths. First, it provides one of the few qualitative assessments of competency-based equipment user training for bubble CPAP implementation in Tanzania, addressing an important evidence gap on how training influences healthcare providers’ competence, clinical practice, and equipment utilization in routine neonatal care. Second, the study included healthcare providers from different professional cadres, including nurses and doctors, across three regional referral hospitals implementing the NEST360 programme. This diversity of participants enabled triangulation of perspectives and provided a comprehensive understanding of training experiences across different clinical roles and settings. Third, the use of a phenomenological qualitative design allowed participants to describe their lived experiences, generating rich contextual evidence that would not have been captured through quantitative approaches alone. These methodological strengths increase the credibility, dependability, and applicability of the findings for informing neonatal training programmes and implementation of medical technologies in similar low-resource settings.

### Limitations of the study

This study has several limitations that should be considered when interpreting the findings. First, the study relied on healthcare providers’ self-reported experiences and perceptions of competency following the training. Consequently, responses may have been influenced by social desirability or recall bias, particularly because participants had undergone competency-based training through the NEST360 programme. Nevertheless, the inclusion of participants from different professional cadres and health facilities enhanced the credibility of the findings by capturing diverse perspectives.

Second, the study was conducted in three regional referral hospitals implementing the NEST360 programme in Dar es Salaam. These facilities may have greater access to equipment, mentorship, and technical support than many lower-level health facilities in Tanzania. Therefore, the findings may not be fully transferable to primary healthcare facilities or hospitals operating under different resource constraints. However, providing a detailed description of the study context enables readers to assess the applicability of the findings to similar settings.

## Conclusion

This study demonstrates that competency-based equipment user training substantially improved healthcare providers’ competence, confidence, and clinical decision-making in the utilization of bubble CPAP for newborn care. The training also strengthened providers’ ability to perform routine equipment maintenance, troubleshoot minor faults, and collaborate effectively with biomedical technicians, contributing to more efficient use of CPAP within neonatal units. Participants further described improvements in clinical practice, reduced unnecessary referrals, and perceived improvements in newborn care.

These findings suggest that competency-based equipment user training is an important component of successful implementation of neonatal technologies in resource-limited settings. Integrating practical, competency-based training into routine newborn care programmes may strengthen healthcare provider capacity and support the effective utilization of life-saving technologies for small and sick newborns.

## Supporting information

Figure 1: Intervention framework

## Acknowledgement

The authors would like to acknowledge the Ifakara Health Institute (IHI), through the NEST360 program, for its valuable institutional and programmatic support throughout this work. We also extend our sincere appreciation to Amana Regional Referral Hospital, Temeke Regional Referral Hospital, and Mwananyamala Regional Referral Hospital for their cooperation and support during the conduct of the study. We gratefully acknowledge the leadership, commitment, and collaboration of the Regional Health Management Teams (RHMTs), and the respective health facility leadership and healthcare providers (respondents). Their dedication and continued support were instrumental in facilitating the successful implementation of this study. Special appreciation is extended to the Muhimbili University of Health and Allied Sciences (MUHAS) for the academic and institutional support provided throughout the Master of Public Health (MPH) training of JM, during which this research was undertaken.

## Data availability statement

No personal identifiable information was collected, and in some cases, interviews were de-identified at the point of transcription. Interested researchers may seek the access approval from the Muhimbili University of Health and Allied Science Ethics Committee via the secretariat at

